# Early deviations from normative brain morphology and cortical microstructure in schizophrenia spectrum disorders

**DOI:** 10.64898/2026.01.09.26343760

**Authors:** Claudio Alemán-Morillo, Natalia García-San-Martín, Richard AI Bethlehem, Patricia Segura, Chloe Gomez, Alessia Pasquini, Pablo Salguero-Quiros, Lena Dorfschmidt, Manuel Muñoz-Caracuel, Rosa Ayesa-Arriola, Javier Vázquez-Bourgon, John Suckling, Miguel Ruiz-Veguilla, Benedicto Crespo-Facorro, Rafael Romero-García

## Abstract

In schizophrenia spectrum disorders (SSD), structural alterations of the gray matter (GM) and white matter (WM) have been widely described. However, the complex interplay between early disease-related changes and ongoing brain maturation challenges our ability to identify early biomarkers. In this study, we investigated structural abnormalities and their association with symptoms in a drug-naïve or minimally medicated sample comprising 113 patients with SSD and 112 neurotypical controls. Specifically, we derived centile scores using normative modelling from cortical thickness (CT), and subcortical volumes derived from structural MRI images, and diffusion tensor imaging-derived (DTI) WM tract fractional anisotropy (FA). In addition, we derived raw cortical mean diffusivity (cMD) metrics from DTI. Compared to controls, SSD participants showed reduced CT centiles, ventricular enlargement, and subcortical centile reductions in the hippocampus, thalamus, amygdala, and nucleus accumbens. SSD was also associated with widespread increases in cMD. We also explored associations among these structural markers, identifying significant relationships between CT centiles and raw cMD, as well as between subcortical centiles and FA tract-based centiles. No interaction with SSD diagnosis was found. Furthermore, positive symptoms correlated negatively with CT and FA tract-based centiles, and showed widespread positive associations with raw cMD, whereas negative symptoms showed no associations. These findings underscore multimodal brain abnormalities that originate early in the course of SSD and their distinct associations with symptoms. Our results support the potential of both normative modelling and diffusion imaging markers to identify individualized early brain changes in SSD Among these, cMD emerges as a potential marker of SSD-related microstructural alterations.

## INTRODUCTION

Schizophrenia spectrum disorders (SSD) comprise a range of clinically heterogeneous conditions, from schizophrenia to brief psychotic disorders, that share core psychotic symptoms and partially overlapping neurobiological alterations despite differences in severity and illness duration, posing important challenges for their investigation and characterization [1–7]. Structural MRI studies have consistently revealed gray matter (GM) abnormalities in SSD individuals. Widespread reductions in cortical thickness (CT) have been consistently reported [8, 9], particularly in frontal and temporal regions [10–12], and, in some cases, in the insula and cingulate cortex [13]. Reductions in subcortical volumes [14, 15] and ventricular enlargement have also been extensively described in schizophrenia [16], with these alterations being associated with poorer functional outcomes and greater severity of negative symptoms (8). Interestingly, studies have also observed volume increases in specific regions such pallidum and caudate [17], highlighting the neuroanatomical variability of this condition.

Interindividual variability of brain structure is strongly influenced by brain maturation, with age and sex being associated with differentiated regional trajectories [18]. To address this problem, normative modeling provides a useful framework for identifying individualized deviations in a measured characteristic from its expected value for the same age and sex in healthy individuals. These differences are expressed in centiles ranging from 0 to 1, with the 0.5 centile assigned to the mean value expected for that age and sex in healthy individuals. Greater deviations from this value indicate a greater abnormality. This approach is especially valuable in SSD, a condition marked by high neurodevelopmental and clinical heterogeneity, which limits the validity of traditional comparisons between groups [19].

Large-scale initiatives, such as those conducted by the ENIGMA Clinical High Risk for Psychosis Working Group, have identified temporal lobe abnormalities in individuals who later transitioned to psychosis [20]. The global CT lifespan trajectories of individuals with psychosis have been described as lower than the expected trajectory of neurotypical individuals, both at the first episode and during chronic stages [21]. These deviations have been associated with clinical outcomes, indicating that reduced CT correlated with greater severity of negative symptoms, particularly in the parietal and temporal regions [22]. Longitudinal research emphasizes the clinical relevance of these deviations, where anomalous reductions in CT over time have been associated with symptom improvement and better cognitive outcomes, particularly among individuals exposed to lower levels of antipsychotic medication [23, 24]. Moreover, deviations in specific cortical regions, such as the superior temporal gyrus and Broca’s area, have been demonstrated to be useful for predicting long-term outcomes [25]. Notably, normative deviations have shown superior predictive value over raw volumetric measures for treatment response, emphasizing the clinical relevance of individual-level normative approaches [24, 26].

In addition to GM abnormalities, microstructural alterations are increasingly recognized as a potential feature in the diagnosis of degenerative and psychiatric diseases. DTI characterizes the anisotropic diffusion of water molecules within tissue, with derived metrics such as Fractional Anisotropy (FA), quantifying the degree of diffusion directionality and providing an indirect measure of white matter integrity. Complementary, Mean Diffusivity (MD) represents the average rate of water diffusion, serving as an indicator of tissue deterioration and a proxy for the loss of water-containing structures. Building on this, cortical MD (cMD) estimates water mobility within the cortical ribboning, reflecting the disruption of microstructural barriers such as myelin or cell membranes that normally restrict the motion of water molecules [27, 28]). Emerging evidence indicates that cortical diffusion MRI metrics capture biologically meaningful features of GM microstructure that extend beyond traditional morphometric measures. Histological findings suggest that cortical diffusion properties are shaped by neurite organization, while recent PET-MRI studies demonstrate that mean diffusivity is closely related to synaptic density in healthy individuals [29]. Together, these findings support the use of diffusion-derived cortical measures as potential in vivo markers of cytoarchitectural and synaptic integrity [30–32]. The extent to which these relationships are altered in SSD, however, remains poorly understood.

A growing body of literature has reported reductions in FA within Deep White Matter (DWM) tracts both in individuals with first episode psychosis and chronic schizophrenia [33]. In adolescents with early-onset psychosis, microstructural abnormalities have also been observed in regions such as the right posterior cingulate gyrus, right superior and posterior corona radiata, and parahippocampal gyrus [34]. Increased MD and decreased FA in the superior and inferior longitudinal fasciculus have been associated with disorganization symptom severity [33]. Within the cortex, cMD has emerged as a sensitive biomarker for detecting microstructural cortical damage, even in the absence of atrophy. Its ability to capture early alterations in neurodegenerative and psychiatric disorders, such as Alzheimer’s [27, 35], Parkinson’s [36, 37], and Huntington’s disease [38], along with treatment-resistant schizophrenia [39], makes it increasingly valuable for early diagnosis. Moreover, cMD has been proposed as a more specific marker of neuronal loss or deterioration than CT [39, 40]. In this context, increased isometric diffusivity has been associated with poorer social functioning and greater clinical symptom severity in SSD, even in the absence of alterations in cortical volume or FA [41]. These findings highlight cMD as a promising, yet underexplored, biomarker of early microstructural disruption in psychosis.

In the present study, we investigate individual-level alterations of raw cMD, together with deviations in CT, subcortical volumes, and FA tracts derived from normative models in a sample of minimally medicated SSD individuals experiencing a first episode of psychosis. Additionally, we examine how these neuroanatomical deviations relate to the severity of positive and negative symptoms. We hypothesized that SSD individuals would exhibit concurrent deviations in cortical structural and microstructural measures compared with controls, reflected by alterations in normative morphometric centiles and raw cortical cMD values. Furthermore, we expected these abnormalities to be associated with clinical symptom severity and to show spatial associations across cortical regions, reflecting both distinct and shared disruptions in the underlying cytoarchitecture.

## Methods

### Subjects

Participant recruitment was conducted through the Program for Attention to the Initial Phases of Psychoses (PAFIP), established in 2001 at Marqués de Valdecilla University Hospital [42]. Individuals in the clinical group met the Diagnostic and Statistical Manual of Mental Disorders (DSM) IV criteria for SSD. Individuals with SSD were antipsychotic-naïve, with the exception of 14 participants who had received antipsychotic treatment prior to study inclusion; in all cases, exposure was limited to less than 20 days (Table S1). Sample comprised 112 neurotypical controls (males = 68; mean age = 29.73 ± 7.42 years) and 113 SSD individuals (males = 74; mean age = 29.82 ± 8.69 years) with the following distribution of DSM-IV diagnoses: schizophrenia (n=50), schizophreniform disorder (n=33), brief psychotic disorder (n=23), and psychosis not otherwise specified (n=7). Diagnoses were confirmed after a six month follow-up. Although the sample comprised different diagnoses within the schizophrenia spectrum, these conditions were examined under the broader SSD framework, as the present study aimed to investigate shared core psychopathological mechanisms that have been consistently reported across spectrum disorders in previous literature [43–45]. Additionally, sensitivity analyses were conducted to evaluate the potential influence of diagnostic subgroup composition by repeating the analyses separately in participants with schizophrenia and those with brief psychotic or schizophreniform disorder.

This sample represents a subset of a larger longitudinal cohort from the PAFIP program [24] for which DTI data are available. A detailed list of inclusion and exclusion criteria is provided in the Supplementary Methods. A comprehensive summary of the demographic and clinical characteristics of the included sample, stratified by sex and diagnosis is provided in the Supplemental Material (Table S2). The study followed the Declaration of Helsinki and was approved by the CEIC-Cantabria Ethics Committee (clinical trial numbers NCT0235832 and NCT02534363) and the University of Seville Ethics Committee (SICEIA 2024-2534). Informed consent was obtained from all subjects involved in the study.

### MRI T1 acquisition

Structural MRI data (3D T1-weighted) were acquired using a 3T Philips Medical Systems MRI scanner (Achieva, Best, The Netherlands) equipped with an 8-channel head coil with the following acquisition parameters: TE = 3.7ms, TR=8.2ms, flip angle=8°, acquisition matrix=256×256, voxel size=0.94×0.94×1 mm and 160 contiguous slices.

All T1w images underwent quality assurance (Q/A) protocol. Firstly, images were visually inspected by trained raters and were rated as “pass”, “borderline”, or “fail”. Images were considered to have “passed” Q/A, when they were of good quality with no motion artefacts, whereas “borderline” images had some motion artifacts but with enough quality to be used for subsequent analysis of structural estimates. Images that “failed” had blurring, ghosting, or severe ringing artifacts and thus were excluded from all subsequent analysis. Of the 228 original structural images, 226 T1w passed visual inspection, with only two rated as borderline. The Euler number of each scan was computed and included in subsequent analyses to account for image quality [46].

T1w images were processed with FreeSurfer v7.4.1 to extract regional volumes and CT for the 68 regions of the Desikan–Killiany atlas, as well as volumes for eight subcortical structures obtained using the automated FreeSurfer subcortical segmentation (aseg) procedure, for which normative models were available [47, 48]: thalamus, pallidum, nucleus accumbens, hippocampus, caudate, amygdala, and the third and fourth ventricles.

### DTI acquisition

Diffusion Tensor Images (DTI) were collected during the same scanning session using TR = 9577 ms, TE = 77 ms, flip angle = 90°, 64 diffusion directions (b = 1300 s/mm²), one b=0 image and isotropic resolution of 2 mm³. DTI images underwent ENIGMA-DTI visual quality assurance (Q/A) protocol (https://github.com/ENIGMA-git/ENIGMA-DTI-TBSS-Protocol). Two scans were excluded based on visual inspection, and one additional scan was removed as an outlier (mean ±5 standard deviations). As a result, 225 subjects with both acceptable structural T1w and DTI images were retained for further analysis.

### Tract-Based Spatial Statistics (TBSS)

TBSS processing were performed following standardized protocols provided by the ENIGMA-DTI consortium, available at ENIGMA-DTI website (https://enigma.ini.usc.edu/protocols/dti-protocols/) and the ENIGMA GitHub repository (https://github.com/ENIGMA-git/ENIGMA-DTI-TBSS-Protocol). These protocols use a standardized white matter skeleton as a mask for tract extraction. The average fractional anisotropy (FA) value was computed across the entire WM skeleton obtained. Regional FA measures were extracted from 24 bilateral or mid-sagittal regions of interest (ROIs) defined by the Johns Hopkins University (JHU) ICBM-DTI-81 WM atlas [49]. These regions included the corpus callosum CC (body, genu, and splenium; BCC, GCC, SCC), the cingulum bundle (cingulate gyrus and parahippocampal parts; CGC, CGH), the corona radiata CR (anterior, posterior, and superior portions; ACR, PCR, SCR), the corticospinal tract (CST), the external capsule (EC), the fornix and stria terminalis (FX, FXST), the internal capsule IC (anterior, posterior, and retrolenticular limbs;ALIC, PLIC, RLIC), the inferior fronto-occipital fasciculus (IFO), the uncinate fasciculus (UNC), the posterior thalamic radiation (PTR), the superior fronto-occipital fasciculus (SFO), the superior longitudinal fasciculus (SLF), and the sagittal stratum (SS).

### Cortical mean diffusivity (cMD)

To evaluate the microstructural integrity of the cortical ribbon, raw cortical MD was estimated by mapping the DTI into their T1 surface space using Freesurfer’s bbregister. Cortical surfaces were mapped using AFNI SUMA tools. MD maps were projected from the cortical volume onto the cortical (pial) surface using the connectome Workbench command -volume-to-surface-mapping’ in “myelin-style” mode (https://ww.humanconnectome.org/). This projection samples the mean MD across the cortical depth. The resulting projected values were mapped back to each subject’s native surface and were parcellated according to the Desikan-Killiany atlas.

### Normative modelling for GM and WM features

CT and subcortical volumes were referenced against normative data derived from a large neurotypical sample of 138,000 individuals, as described in Bethlehem et al. (2022) [50] and Dorfschmidt et al. (2025) [48]. Using an out-of-sample normative modelling approach (https://github.com/ucam-department-of-psychiatry/Lifespan), we computed centile scores for each individual by comparing their brain measures to a sex- and age-matched reference sample, as well as accounting for site-specific effects. Centiles ranged between 0 and 1, with values below 0.5 indicating volumes lower than the age- and sex-adjusted median, whereas values above 0.5 indicate higher volumes than the median. Despite these corrections, recent studies have shown that age and sex effects can account for a substantial proportion of the variance in case–control differences and in associations between brain structure and symptom severity [51]. Accordingly, age and sex were also included as covariates in the regression models. Centiles for the lateral ventricles could not be computed because normative models for these structures were not available at the time of the analyses.

Similarly, centile values for FA across WM tracts (referred here as tract centiles) were computed using the data and normative modelling approach described by Zhu al, 2024 [52]. FA centiles were estimated using the lifespan reference curves included in the ’eHarmonize’ Python package (https://github.com/ahzhu/eharmonize). Thus, we quantified individual deviation from normative FA in major tracts. As no normative models currently exist for cMD, these analyses were conducted using raw cMD values.

### Clinical assessments

Clinical assessments were obtained from participants diagnosed with SSD and included: (i) Clinical Global Impressions scale (CGI), which provides a comprehensive summary of overall clinical status based on illness severity and global improvement; (ii) Disability Assessment Scale (DAS), which evaluates functional impairment and disability levels; (iii) Brief Psychiatric Rating Scale (BPRS, 24 items); (iv) Scale for the Assessment of Positive Symptoms (SAPS, 30 items); and (v) Scale for the Assessment of Negative Symptoms (SANS, 21 items). In addition, average cognitive functioning was computed across seven domains: (1) verbal memory: Rey Auditory Verbal Learning Test (RAVLT) long-term recall score; (2) visual memory: Rey Complex Figure test (RCFT) long-term recall score; (3) motor dexterity: grooved pegboard (GP), time to complete with dominant hand; (4) executive functions: Trail Making Test part B (TMTb); (5) working memory: WAIS III-Backward Digits (BD) total score; (6) speed of processing: WAIS III-Digit Symbol (DS) standard total score; (7) attention: Continuous Performance Test Degraded-Stimulus (CPT-DS), the total number of correct responses. All assessments were conducted by experienced psychiatrists.

### Association between SSD diagnosis and structural features

To assess the effect of SSD diagnosis on morphometric and microstructural features, multivariate regressions were performed separately for each brain region or tract:

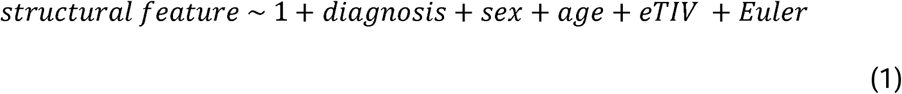

where the dependent variable corresponded to one of the four features analysed in the study (CT centiles, subcortical centiles, tracts centiles, or raw cMD), and *diagnosis* was a binary variable (control or SSD). Given the established association between volume and thickness with both intracranial volume and head movement, estimated Total Intracranial Volume (eTIV) and *Euler* number were included as covariates in those regressions involving CT and subcortical volumes. In contrast, these variables were not included for FA and cMD regressions, as these are microstructural features are not influenced by overall brain size (FA, r=0.09, p=0.15; cMD r=0.10, p=0.86) or Euler number (FA, r=0.01, p=0.39; cMD r=0.11, p=0.11) To control for multiple comparisons, *p*-values obtained from the regional and tracts regressions were corrected using False Discovery Rate (FDR) using 0.05 as threshold. Additionally, sensitivity analyses excluding previously medicated SSD participants were performed to evaluate the potential influence of antipsychotic exposure.

### Interrelationships among GM and microstructural features

We next conducted multivariate linear regressions to investigate whether GM morphometric features (CT and subcortical volume centiles) were associated with microstructural features (FA centiles and raw cMD), and whether these associations were moderated by SSD diagnosis:

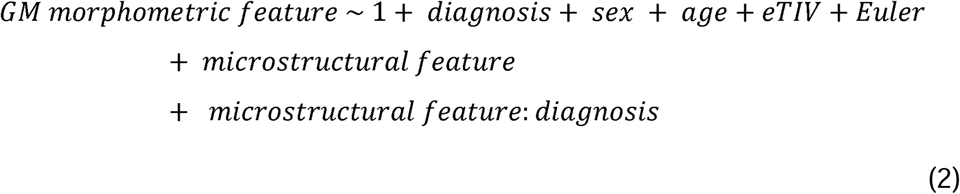

Accordingly, three separate analyses were conducted: (i) each regional CT centile was compared with the corresponding regional raw cMD; (ii) since there is no direct spatial correspondence between subcortical regions and white matter tracts, comparisons were based on global averages, with subcortical centiles compared to the mean FA tract centile; and (iii) conversely, each FA tract centile was compared to the mean subcortical centile. All p values were corrected using False Discovery Rate (FDR) using 0.05 as threshold.

### Structural features and clinical variables

First, we examined the associations between the patients’ global structural features and a range of clinical variables, including the CGI, DAS, overall cognitive functioning, and symptom severity as assessed by the BPRS, SAPS, and SANS scales. Additionally, characteristics of the first episode were also considered, including whether hospitalization occurred (yes/no), its duration when applicable, and the presence of a family history of psychosis.

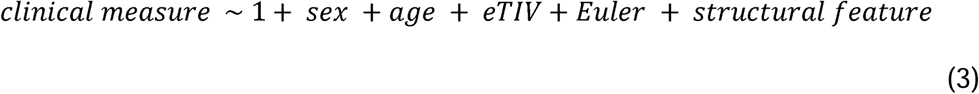

*Structural features* included CT, subcortical, and tract centiles, along with raw cMD. eTIV and *Euler* number were included only in models involving GM morphometric features due to the aforementioned reasons. Additionally, analyses were conducted separately for each symptom scale and each regional structural feature. All p values were FDR corrected using 0.05 as threshold.

As a sensitivity analysis, these models were repeated using raw structural values (CT, Subcortical volumes and FA tracts) instead of normative metrics.

## RESULTS

### Impact of SSD diagnosis in structural features

We first explored the impact of SSD diagnosis and the included covariates on the four structural features considered: (i) CT centiles, (ii) subcortical volumetric centiles, (iii) FA tracts centiles, and (iv) raw cMD.

Mean CT centiles were reduced in individuals with SSD (T=-2.52, p=0.01), with females showing lower values than males (T=-3.00, p<10^-2^) and a positive correlation with age (T=2.41, p=0.02). At the regional level, widespread significant reductions of CT centiles were found at parietal (T<-2.90, p<0.04) and cingulate (T<-2.94, p=0.04) cortices (Fig. 1A). Subcortical volumetric centiles revealed that SSD had significantly larger third (T=4.13, p<10^-3^) and fourth (T=3.66, p<10^-2^) ventricles (Fig. 1B). Additionally, reduced centiles were observed in several subcortical GM structures, including the left accumbens (T=-2.90, p=0.01), bilateral thalamus (right T=-2.48, p=0.03; left T=-2.52, p=0.03), right amygdala (T=-2.59, p=0.03) and bilateral hippocampus (right: T=-2.87, p=0.01; left: T=-2.99, p=0.01). No significant differences in tracts centiles survived FDR correction (Fig. 1C). Nonetheless, the anterior corona radiata (T=-2.70, p=0.09) and genu of the corpus callosum (T=-3.06, p=0.06) showed trends toward significance. Individuals with SSD showed widespread raw cMD increased values in frontal (T>2.64, p<0.02), cingulate (T>2.52, p<0.02), occipital (T>2.31, p<0.03), parietal (T>2.20, p<0.04), and temporal (T>2.15, p<0.04) regions (Fig. 1D). In a sensitivity analysis, using raw data instead of normative data in the models, the only difference observed was a significant relationship between diagnosis and decreases in FA in the GCC, FX, EC and ACR tracts (Fig. S1). Full details on these regional effect sizes are reported in the Supplementary Data.

**Figure 1.**
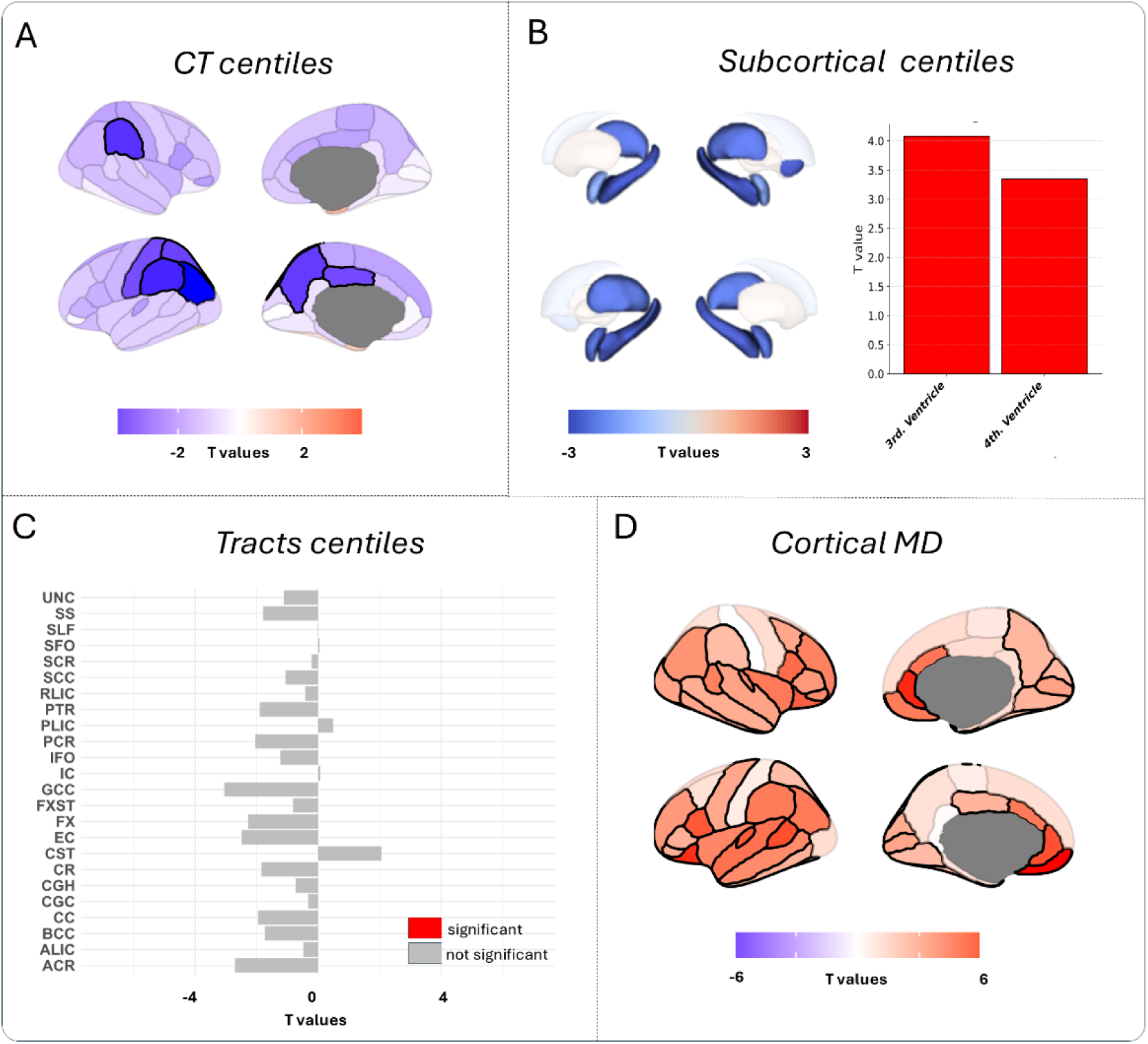
Effect sizes (T-values) reflecting the contribution of SSD diagnosis, after considering sex, age, eTIV, and Euler number, across four structural features: (A) CT centiles, (B) subcortical volumetric centiles, (C) FA tract centiles, and (D) raw cMD. Colour scales range from blue (negative T-values indicating reduced structural features in SSD compared to control) to red (positive T-values indicating increased structural features in SSD compared to control). Regions that did not survive FDR correction are shown with desaturated semi-transparent shading, whereas significant regions are displayed in solid colours. Significant tracts and ventricles are shown in solid red.

Sensitivity analyses excluding previously medicated participants revealed similar regional effect sizes (Fig. S2). Additional subgroup analyses separating schizophrenia from brief psychotic disorder and schizophreniform disorder also revealed, despite expected differences in effect size magnitude between subgroups, consistent spatial patterns across regions (Fig. S3).

### Normative associations between GM morphometric and microstructural features

To investigate both normative and atypical associations between GM morphometric and microstructural features, we examined the correlations among centile-based metrics and raw cMD. CT centiles were significantly associated with raw cMD (T=-4.08, p<0.01), while subcortical volumetric centiles were correlated with FA tracts centiles (T=5.04, p<0.01). However, across all models, the “diagnosis × structural feature” interaction term was non-significant, indicating no detectable interaction with SSD. Overall, these results reveal that individuals with higher CT centiles exhibited lower raw cMD, whereas those with higher subcortical centiles had increased FA tracts centiles. However, these relationships were not influenced by the clinical condition.

Based on these global associations, we conducted a regional analysis to identify localised effects. CT centiles of the cingulate cortex showed a positive association with raw cMD (Fig. 2A), reaching significance in bilateral isthmus cingulate (T>2.91, p<0.03) and left posterior cingulate (T=2.88, p=0.03). In contrast, negative associations between CT centiles and raw cMD were found in right paracentral (T=-4.48, p<0.01), bilateral precuneus (T<-3.45, p<0.01), bilateral superior parietal (T<-3.59, p<0.01), and left temporal pole (T=-3.15, p=0.01). Subcortical volumetric centiles were associated with average FA tracts centiles for the left thalamus (T=3.13, p=0.04) (Fig. 2B). Conversely, several tract centiles were associated with average subcortical volumetric centiles, including the genu of the corpus callosum (T=3.11, p=0.01), inferior fronto-occipital fasciculus (T=2.72, p=0.03), fornix-stria terminalis (T=4.00, p<0.01), cingulum (T=3.37, p<0.01), corpus callosum (T=3.11, p=0.01), and its body (T=4.04, p<0.01) (Fig. 2C). Consistent with the previous global findings, no significant interaction with SSD diagnosis was observed in any of the regional models.

**Figure 2.**
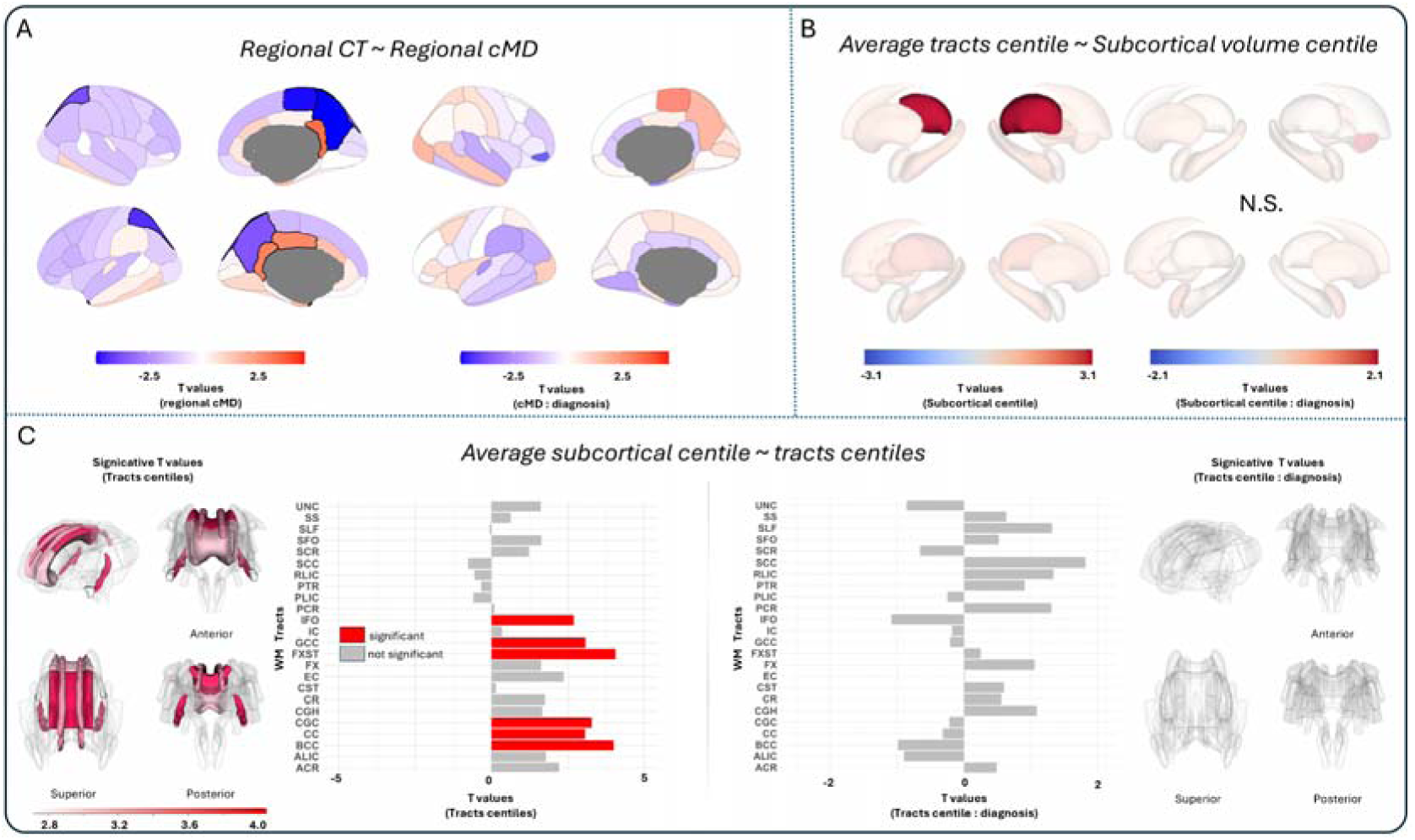
T-values reflecting the associations between GM morphometric and microstructural features: (A) regional CT centiles and raw cMD, including diagnosis interactions; (B) subcortical volumetric centiles and mean FA tract centiles, including diagnosis interactions; and (C) FA tract centiles and mean subcortical GM volume. Colour scales range from blue (negative T-values indicating anti-correlated structural features) to red (positive T-values indicating positively correlated structural features). Regions that did not survive FDR correction are shown with desaturated semi-transparent shading, whereas significant regions are displayed in solid colours. Significant tracts and ventricles are shown in solid red.

### Relationship between structural features and clinical symptomatology

We finally explored whether the four structural features were associated with clinical status and symptomatology.

Mean raw cMD was significantly and positively associated with hospitalization (T=2.7, p=0.02), CGI (T=3.51, p<0.01) and SAPS scores (T=3.71, p< 0.01) (**Table 1**). In contrast, mean CT centile showed a significant negative correlation with days of hospitalization (T=–2.63, p=0.04) and with CGI (T=-2.99, p=0.03). Finally, the mean centiles of the third and fourth ventricles were significantly and positively correlated with SAPS (T=3.14, p=0.02). No other associations remained significant after FDR correction.

**Table 1.**
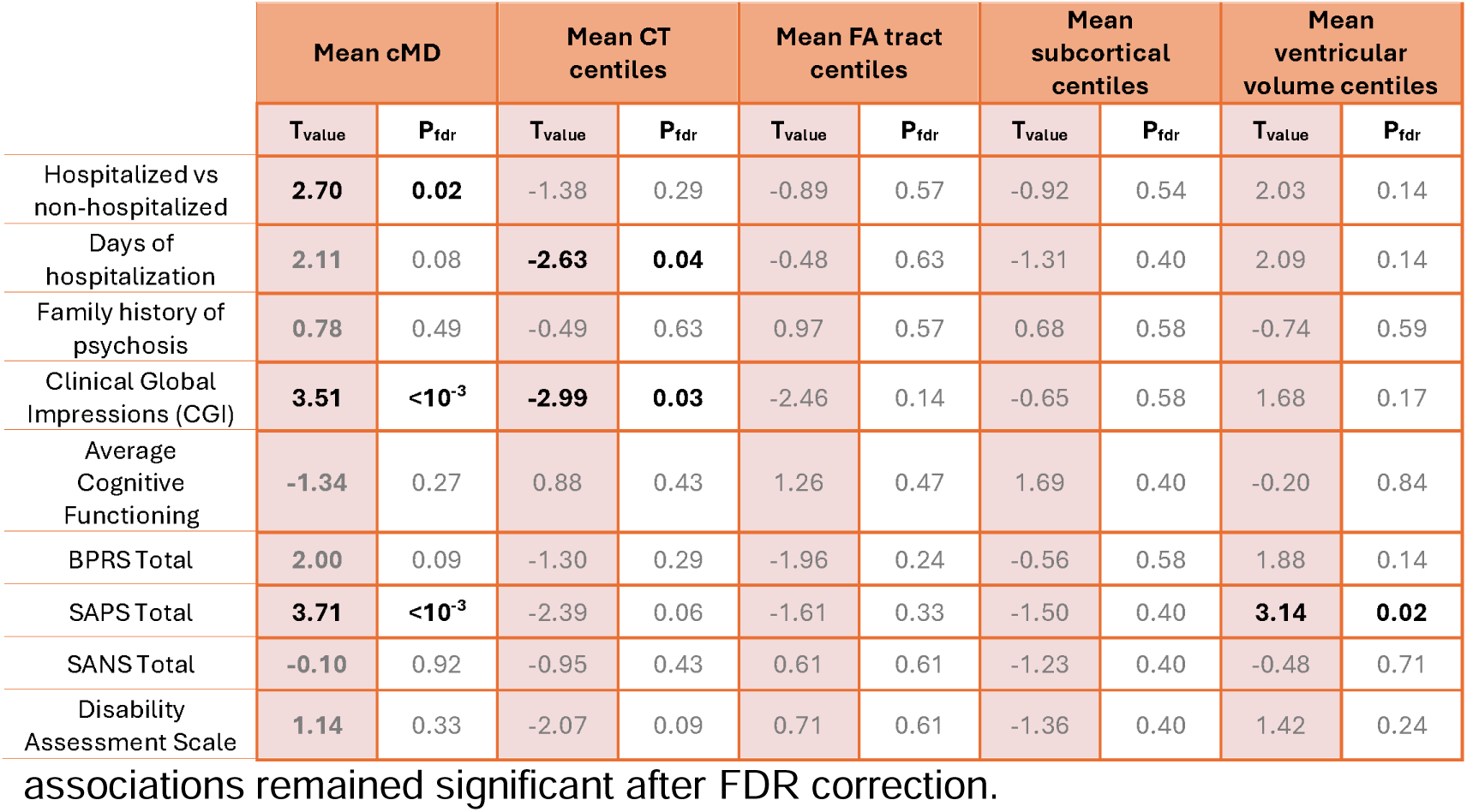
Summary of regression analyses relating mean structural features to clinical characteristics assessed at the first episode of psychosis.

In the regional analyses (Fig. 3), correlations between BPRS scores and both GM morphometric features and cMD did not survive FDR correction. BPRS was significantly correlated with the FA centile of two WM tracts: sagittal stratum (T=-3.00, p=0.04) and the posterior thalamic radiation (T=-3.20, p=0.04). The association between CT centiles and SAPS survived the FDR threshold for the right superior temporal gyrus (T=-3.54, p=0.03), supramarginal gyrus (T=-3.28, p=0.03), and frontal pole of the right (T=-3.33, p=0.03). Correlations between SAPS and subcortical centiles did not survive corrections, except for the third ventricle, which exhibited a positive association (T=3.59, p=0.01). A widespread positive correlation between SAPS and cMD was found (cingulate T>2.72, p<0.02; frontal T>2.31, p<0.04; insula T>3.49, p<0.01; occipital T>2.8, p<0.015; parietal T>2.48, p<0.03; temporal T>3.32, p<0.01). None of the negative associations between SAPS and regional tract centiles remained significant after correction. SANS did not show significant associations with any of the features examined. The sensitivity analysis using raw structural data instead of normative measures yielded largely similar results. The main differences were the absence of a significant association between ventricular measures and SAPS, and the emergence of strong associations between BPRS scores and FA in several white matter tracts (SS, RLIC, PTR, PCR, IC, EC, CR, CGH and ALIC) (Fig. S4).

**Figure 3.**
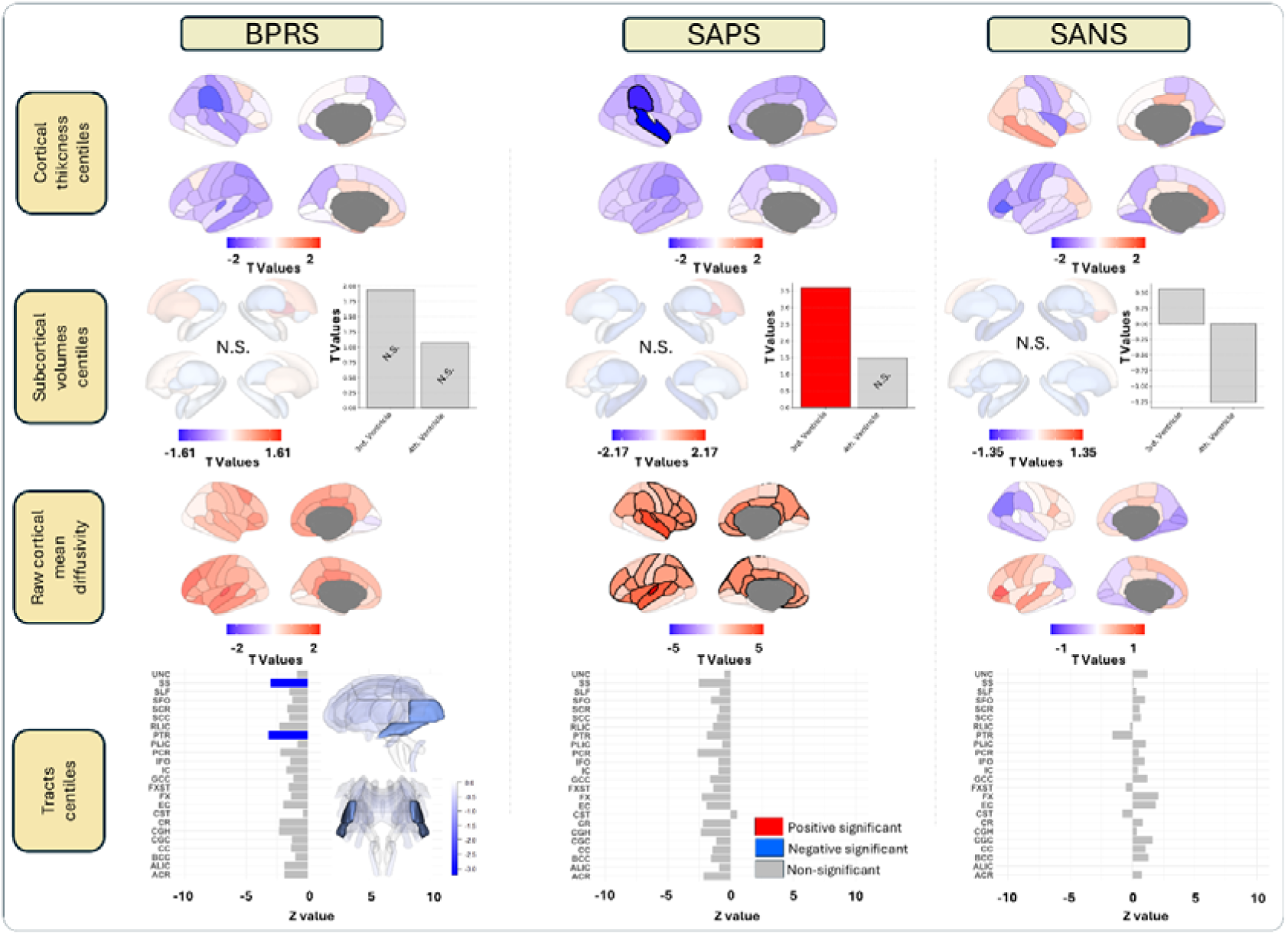
T-values reflecting the associations between symptom scores (BPRS, SAPS, and SANS) and regional structural features, including CT centiles, subcortical volumetric centiles, raw cMD, and FA tract centiles. Colour scales range from blue (negative T-values indicating structural features anti-correlated with symptoms) to red (positive T-values indicating structural features positively correlated with symptoms). Regions that did not survive FDR correction are shown with desaturated semi-transparent shading, whereas significant regions are displayed in solid colours. Significant tracts and ventricles are shown in solid colour depending on whether they are positively or negatively correlated.

## DISCUSSION

In our study we found that deviations from normative brain macrostructure, as well as less explored microstructural features in psychosis, such as cMD, are significantly associated with diagnosis and clinical symptoms in the early stages of psychosis. Consistent with prior studies [11], we observed generalized reductions in CT in SSD patients compared to neurotypical controls. These findings align with normative [22] and non-normative [53] studies reporting CT and volumetric alterations in frontal, parietal, and cingulate regions. Subcortical volume reductions were also consistent with previous findings showing decreased normative volumes in the amygdala, thalamus, and hippocampus [26, 54]. Enlargement of the ventricles, including the third ventricle aligns with longstanding evidence linking ventricular expansion to psychosis [55–57]. This enlargement had been mainly associated with poorer outcomes in general [16] and negative symptomatology [55] in SSD.

Similar to the patterns observed for CT centiles, raw cMD showed a generalized and widespread increase in the SSD group, reflecting greater water mobility within the cortex. This may indicate cortical tissue loss or microstructural disruption of the cortical architecture that typically restricts water diffusion, such as cell membranes or the amount of myelin [27, 39, 58]. In this line, previous studies have consistently reported significant increases in cMD in temporal, parietal, frontal, insular, and occipital cortices [59–61], highlighting the broad spatial extent of cortical microstructural abnormalities associated with this condition.

The widespread reductions in GM features observed in SSD patients contrasted with the limited effect sizes seen in FA tract centiles. Although these WM alterations were not significant, we observed a reduced general pattern, where, consistent with previous findings, the most pronounced effects were found in the anterior corona radiata and the genus of the corpus callosum [62]. The lack of statistically significant findings does not necessarily imply the absence of FA reductions in the tracts. Rather, it may reflect the presence of heterogeneous patterns of FA alterations across individuals with SSD according to previous studies using normative models that highlight individual variability in WM alterations [63].

As expected, the four structural features were not independent. We observed positive correlations between CT centiles and raw cMD in cingulate areas, and negative correlations in parietal regions for controls. These regional patterns are consistent with prior studies that suggest that the relationship between CT and microstructure varies by cortical functional architecture, reflecting distinct coupling in polymodal versus unimodal regions [64]. Moreover, we identified a positive association between thalamic volumetric centile and mean FA, supporting previous evidence linking thalamic volume to the integrity of thalamo-cortical WM projections, particularly in anterior pathways [65]. This relationship may underlie the broader positive association that we observed between multiple tracts and overall subcortical volume. Interestingly, none of these associations were moderated by the diagnosis by feature interaction term, indicating that the clinical diagnosis does not influence the relationships among structural features.

Regarding the characteristics of the first psychotic episode, mean global CT was associated with days of hospitalization and CGI scores. Regional analyses revealed that reduced temporal and parietal CT centiles, as well as increased ventricular centiles, were associated with greater severity of positive symptoms. In particular, the superior temporal gyrus has been consistently implicated, with several studies reporting associations between positive symptom severity and cortical thinning in schizophrenia, particularly in the temporal region [66–68]. It has been hypothesized that these alterations of the temporal regions may play a key role in cognitive impairment, as well as in disorganized behaviour or semantic memory dysfunction, with the latter being associated with delusions and disorganized thinking [69]. A normative study in chronic schizophrenia patients also reported associations between frontal and temporal CT with hallucinations, delusions, and depression (44). Similarly, the supramarginal gyrus, located within the inferior parietal lobule, has been associated with language processing and storage [70] and social cognition [71], highlighting its potential role in psychosis (55,56). Similarly, third ventricular enlargement has been associated with negative symptomatology in SSD [72], which, according to previous findings, could be linked to cognitive impairment [57]. However, in our results, third ventricular enlargement was instead associated with positive symptomatology.

Regarding cMD global, it shows a significant relationship with the need for hospitalization and the CGI score. Furthermore, it also shows a relationship with the SAPS score. The regional analyses also revealed that positive symptomatology was extensively associated with increased regional raw cMD values, with the strongest effect observed in the superior temporal cortex. This aligns with prior findings linking elevated cMD in the temporal gyrus to poorer social functioning and higher PANSS scores [41]. Our study showed a widespread increase in raw cMD, suggesting a generalized pattern of microstructural deterioration, consistent with our findings on CT. Although previous studies have reported that increased cMD in the right insular cortex was associated with negative symptoms, supporting the idea that fronto-temporal microcircuit disruption may contribute to schizophrenia pathophysiology [60], we did not find significant associations between raw cMD and negative symptoms in any cortical region. The absence of a relationship with negative symptoms in our sample may be explained by the very low levels of negative symptoms observed in our cohort (SANS = 0.98 ± 1.32). Altogether, these results suggest that CT centiles and raw cMD could represent a particularly sensitive biomarker of clinical status.

Deep White Matter (DWM) tract analyses revealed a negative correlation between FA centiles in the posterior thalamic radiation and sagittal stratum and BPRS scores, a measure of clinical severity in psychosis. The posterior thalamic radiation includes thalamocortical fibers such as the optic radiation, while the sagittal stratum comprises major associative pathways, including the inferior fronto-occipital fasciculus, both of which have been reported as affected in psychosis in previous studies [73, 74] and related to hallucinations [74, 75]. These findings align with the theory of occipital cortical disconnection, which posits that impaired integration of visual input may contribute to psychotic symptomatology [76]. Reduced FA in these tracts may reflect microstructural disruptions within the visual and associative circuitry [77], suggesting that abnormalities in the functioning of this system may serve as early markers of psychosis [78, 79].

The sensitivity analyses using raw data generally showed consistent results to those obtained using normative metrics. Although some specific differences were observed, especially in tracts analyses, these generally followed the same directional patterns as the normative results. This suggests that normative approaches may provide a more conservative framework for establishing brain–behaviour associations, potentially by accounting for relevant non-linear age-and sex-related effects

## LIMITATIONS

Several limitations must be acknowledged. First, the primary inclusion criterion was the diagnosis of SSD, which predominantly included schizophrenia but also other psychotic disorders so we cannot specify the results for each disorder. Second, given the limited effect sizes reported here, larger cohorts may be needed to validate subtler associations or subgroup effects. Third, the cross-sectional design limits our ability to draw conclusions about developmental trajectories. Longitudinal studies are required to characterize the progression of microstructural changes over time and evaluate their potential utility as prognostic markers. Fourth, given the low SANS scores observed in our patient sample, the statistical power to detect associations with negative symptoms was lower than for the other symptom dimensions explored in this study. Fifth, although hospitalization and its duration may reasonably correlate with first-episode severity, they are also influenced by patient-specific factors, clinical features, and institutional practices. Consequently, hospitalization should not be interpreted as a direct measure of severity.

## CONCLUSIONS

In summary, our findings provide converging evidence that early-stage psychosis is characterized by a distinct pattern of structural alterations involving cortical, subcortical, and WM alterations. Raw cMD showed the highest sensitivity to positive symptoms and was associated with key clinical features at the first episode, including hospitalization and overall illness severity. While reductions in CT, ventricular enlargement, and increased cortical diffusivity were prominent, weaker effect sizes were found in DWM. The differential correlations between symptom dimensions and structural features support the presence of distinct neurobiological mechanisms underlying positive, negative, and global psychopathology. These results emphasize the need for multidimensional clinical assessment and suggest that early deviations from normative brain structure, together with alterations in raw cMD measures, may reflect microstructural and macrostructural abnormalities associated with SSD.

## Data and Code availability

All code and non-clinical data used to perform the analyses can be found at https://github.com/NeuroimagingBrainNetworks/cortical_microstructure_in_SSD

## Acknowledgements

We thank Victor Ortiz and Jakob Seidlitz for their contribution.

## Authors contributions

C.A.M. performed data curation, methodological design, data analysis, and drafted the manuscript; N.G.S.M, R.A.B, P.S, C.G, A.P, P.S.Q, L.D, M.M.C, R.A.A, J.V.B, J.S., M.R.V, B.C.F and R.R.G contributed to data acquisition, provided advice on data analysis, and participated in writing and editing the manuscript. R.R.G. also contributed to conceptualization and supervision of the work. All authors approved the submitted version of the manuscript.

