## Supplemental Material for "Early deviations from normative brain morphology and cortical microstructure in schizophrenia spectrum disorders"

Supplementary Material

### Participants

Data for the present study were drawn from a large cohort of individuals with a first episode of psychosis (FEP) in Cantabria, a region in northern Spain with epidemiological representativeness. Participants were enrolled in PAFIP (Programa de Atención a las Fases Iniciales de Psicosis), a multidisciplinary intervention program conducted at the University Hospital Marqués de Valdecilla. Referrals came from inpatient units, emergency services, mental health services, and other healthcare workers across the region.

Inclusion criteria included: age 15–60 years, residence within the catchment area, FEP diagnosis, and either no prior or minimal antipsychotic exposure (less than six weeks). Exclusion criteria were DSM-IV diagnoses of drug or alcohol dependence, intellectual disability, or history of neurological disease or head injury. Diagnoses were confirmed using the Structured Clinical Interview for DSM-IV (SCID-I) conducted by an experienced psychiatrist within 6 months of first episode of psychosis visit.

Sociodemographic and clinical data were obtained from interviews and medical records. Symptom dimensions were evaluated using the Scale for the Assessment of Positive Symptoms (SAPS), the Scale for the Assessment of Negative Symptoms (SANS) and the Brief Psychiatric Rating Scale (BPRS).

Healthy controls were assessed using a condensed version of the Comprehensive Assessment of Symptoms and History, ensuring absence of psychiatric or neurological disorders, significant medical conditions, or substance dependence.

### Supplementary tables

| **Previous Treatment** | **Nº of patients** | **Duration (days)** | **Standard deviation (days)** |
| --- | --- | --- | --- |
| Aripiprazole | 2 | 5.5 | 3.54 |
| Risperidone | 5 | 8.2 | 5.54 |
| Olanzapine | 6 | 7.17 | 10.4 |
| Unknow | 1 | 1 | 0 |
| Total | 14 |  |  |

**Table S1.** Exposure to previous antipsychotic treatment.

|  | Male  Control | Female Control | Male  SSD | Female  SSD |
| --- | --- | --- | --- | --- |
| Age | 29.47 ± 6.52 | 30.12 ± 8.69 | 28.57 ± 8.37 | 32.18 ± 8.91 |
| Age of onset |  |  | 27.78 ± 8.33 | 32.08 ± 8.87 |
| CPZ equivalent |  |  | 159.19 ± 61.28 | 158.59 ± 49.27 |
| SANS Unchanging Facial Expression |  |  | 0.86 ± 1.39 | 0.43 ± 1.04 |
| SANS Decreased Spontaneous Movements |  |  | 0.33 ± 0.98 | 0.05 ± 0.33 |
| SANS Paucity of Expressive Gestures |  |  | 0.17 ± 0.68 | 0.05 ± 0.33 |
| SANS Poor Eye Contact |  |  | 0.39 ± 1.11 | 0.08 ± 0.36 |
| SANS Affective Non responsivity |  |  | 0.25 ± 0.85 | 0.11 ± 0.46 |
| SANS Inappropriate Affect |  |  | 1.3 ± 1.83 | 1.69 ± 2.04 |
| SANS Lack of Vocal Inflections |  |  | 0.34 ± 1.03 | 0.16 ± 0.73 |
| SANS Global Rating of Affective Flattening |  |  | 1.22 ± 1.55 | 0.49 ± 1.05 |
| SANS Poverty of Speech |  |  | 0.45 ± 1.25 | 0.29 ± 0.84 |
| SANS Poverty of Content of Speech |  |  | 0.22 ± 0.86 | 0.03 ± 0.16 |
| SANS Blocking |  |  | 0.32 ± 1.05 | 0.13 ± 0.66 |
| SANS Increased Latency of Response |  |  | 0.46 ± 1.23 | 0.45 ± 1.03 |
| SANS Global Rating of Alogia |  |  | 0.84 ± 1.51 | 0.62 ± 1.25 |
| SANS Grooming and Hygiene |  |  | 0.4 ± 0.96 | 0.21 ± 0.92 |
| SANS Inpersistence at Work or School |  |  | 0.87 ± 1.59 | 0.36 ± 1.2 |
| SANS Physical Anergia |  |  | 0.68 ± 1.31 | 0.21 ± 0.61 |
| SANS Global Rating of Avolition Apathy |  |  | 1.12 ± 1.66 | 0.44 ± 1.23 |
| SANS Recreational Interests and Activities |  |  | 0.9 ± 1.6 | 0.57 ± 1.28 |
| SANS Sexual Activity |  |  | 0.56 ± 1.29 | 0.27 ± 0.9 |
| SANS Ability to Feel Intimacy and Closeness |  |  | 0.79 ± 1.6 | 0.54 ± 1.37 |
| SANS Relationships with Friends and Peers |  |  | 1.35 ± 1.86 | 0.65 ± 1.42 |
| SANS Global Rating of Anhedonia Asociality |  |  | 1.55 ± 1.88 | 0.87 ± 1.54 |
| SANS Social Inattentiveness |  |  | 0.29 ± 0.93 | 0.67 ± 1.44 |
| SANS Inattentiveness During Mental Status Testing |  |  | 1.91 ± 2.08 | 2.03 ± 2.02 |
| SANS Global Rating of Attention |  |  | 2.07 ± 2.09 | 2.08 ± 1.98 |
| SAPS Auditory hallucinations |  |  | 1.68 ± 2.23 | 1.07 ± 1.94 |
| SAPS Voice commenting |  |  | 0.66 ± 1.68 | 0.75 ± 1.69 |
| SAPS Voice conversing |  |  | 0.77 ± 1.8 | 0.5 ± 1.35 |
| SAPS Somatic or tactile hallucinations |  |  | 0.32 ± 1.24 | 0.43 ± 1.32 |
| SAPS Olfactory hallucinations |  |  | 0.04 ± 0.29 | 0.25 ± 0.97 |
| SAPS Visual Hallucinations |  |  | 0.11 ± 0.73 | 0.07 ± 0.26 |
| SAPS Global rating of hallucinations |  |  | 2.73 ± 2.36 | 2.26 ± 2.37 |
| SAPS Persecutory delusions |  |  | 3.87 ± 2.01 | 4.33 ± 1.46 |
| SAPS Delusions of jealousy |  |  | 0.16 ± 0.9 | 0.28 ± 1.18 |
| SAPS Delusions of guilt or sin |  |  | 0 ± 0 | 0 ± 0 |
| SAPS Grandiose delusions |  |  | 0.16 ± 0.73 | 0.56 ± 1.62 |
| SAPS Religious delusions |  |  | 0.39 ± 1.23 | 0 ± 0 |
| SAPS Somatic delusions |  |  | 0.32 ± 1.25 | 0.22 ± 0.73 |
| SAPS Delusions of reference |  |  | 4.71 ± 0.94 | 4.83 ± 0.51 |
| SAPS Delusions of being contrrolled |  |  | 0.48 ± 1.5 | 0.39 ± 1.2 |
| SAPS Delusions of mind reading |  |  | 1 ± 2 | 0.89 ± 1.81 |
| SAPS Thought broadcasting |  |  | 0.84 ± 1.86 | 0.67 ± 1.57 |
| SAPS Thought insertion |  |  | 0.52 ± 1.5 | 0.33 ± 1.19 |
| SAPS Thought withdrawal |  |  | 0.35 ± 1.25 | 0.33 ± 1.19 |
| SAPS Global rating of delusions |  |  | 4.9 ± 0.38 | 4.9 ± 0.38 |
| SAPS Clothing and appearance |  |  | 0.22 ± 0.71 | 0.27 ± 0.7 |
| SAPS Social and sexual behavior |  |  | 3.14 ± 2.09 | 4.23 ± 1.11 |
| SAPS Aggressive and agitated behavior |  |  | 2.53 ± 2.16 | 2.14 ± 2.14 |
| SAPS Repetitive or stereotyped behavior |  |  | 0.33 ± 0.96 | 1.36 ± 1.81 |
| SAPS Global rating of bizzare behavior |  |  | 4.52 ± 1.14 | 4.54 ± 0.94 |
| SAPS Derailment |  |  | 0.07 ± 0.5 | 0.27 ± 0.84 |
| SAPS Tangentiality |  |  | 0.16 ± 0.71 | 0.89 ± 1.65 |
| SAPS Incoherence |  |  | 0.16 ± 0.71 | 1 ± 1.72 |
| SAPS Illogicality |  |  | 0.15 ± 0.78 | 0.97 ± 1.71 |
| SAPS Circumstantiality |  |  | 0.07 ± 0.4 | 0.57 ± 1.19 |
| SAPS Pressure of speech |  |  | 0.13 ± 0.65 | 0.38 ± 1.01 |
| SAPS Distractible speech |  |  | 0.15 ± 0.72 | 0.32 ± 0.94 |
| SAPS Clanging |  |  | 0.02 ± 0.12 | 0.19 ± 0.7 |
| SAPS Global rating of positive formal thought disorder |  |  | 0.6 ± 1.46 | 1.62 ± 2.02 |
| BPRS bprsom |  |  | 2.44 ± 2.11 | 2.62 ± 2.27 |
| BPRS bprans |  |  | 2.85 ± 1.79 | 3.95 ± 1.93 |
| BPRS bprdep |  |  | 2.67 ± 1.76 | 2.54 ± 1.8 |
| BPRS bprcul |  |  | 1.66 ± 1.57 | 2.21 ± 1.85 |
| BPRS bprhos |  |  | 3.38 ± 2.42 | 3.59 ± 2.45 |
| BPRS bprsus |  |  | 6.56 ± 1.36 | 6.67 ± 1.03 |
| BPRS bprpen |  |  | 6.84 ± 0.58 | 6.74 ± 0.75 |
| BPRS bprgra |  |  | 1.22 ± 0.89 | 1.26 ± 1.12 |
| BPRS bpralu |  |  | 4.05 ± 2.84 | 3.51 ± 2.87 |
| BPRS bprori |  |  | 1.05 ± 0.47 | 1.18 ± 0.64 |
| BPRS bprcon |  |  | 1.63 ± 1.61 | 3.05 ± 2.48 |
| BPRS bprexc |  |  | 2.07 ± 2 | 3.36 ± 2.32 |
| BPRS bprlen |  |  | 1.93 ± 1.49 | 1.33 ± 0.96 |
| BPRS bpremb |  |  | 2.29 ± 1.77 | 1.54 ± 1.23 |
| BPRS bprten |  |  | 2.66 ± 1.9 | 3.18 ± 2 |
| BPRS bprpos |  |  | 1.82 ± 1.64 | 2.1 ± 1.9 |
| BPRS bprcoo |  |  | 2.48 ± 1.97 | 3.21 ± 2.33 |
| BPRS bpremo |  |  | 2.71 ± 2.18 | 2.95 ± 2.24 |
| BPRS bprsui |  |  | 1.62 ± 1.55 | 1.23 ± 0.78 |
| BPRS bprcui |  |  | 1.78 ± 1.48 | 1.51 ± 1.21 |
| BPRS bprext |  |  | 6.08 ± 1.49 | 6.28 ± 1.38 |
| BPRS bprani |  |  | 1.15 ± 0.68 | 1.38 ± 1.27 |
| BPRS bpract |  |  | 1.26 ± 0.83 | 1.62 ± 1.43 |
| BPRS bprdis |  |  | 1.41 ± 1.08 | 1.59 ± 1.41 |
| SANS Total |  |  | 1.18 ± 1.45 | 0.6 ± 0.95 |
| SAPS Total |  |  | 3.19 ± 0.78 | 3.33 ± 0.9 |
| BPRS Total |  |  | 63.62 ± 13.61 | 68.59 ± 16.84 |

**Table S1.** Sociodemographic and clinical characteristics of participants with schizophrenia spectrum disorders (SSD) and healthy controls. Symptom values (SAPS, SANS, and BPRS) correspond to standardized scores. All data are presented as mean ± standard deviation.

### Supplementary figures

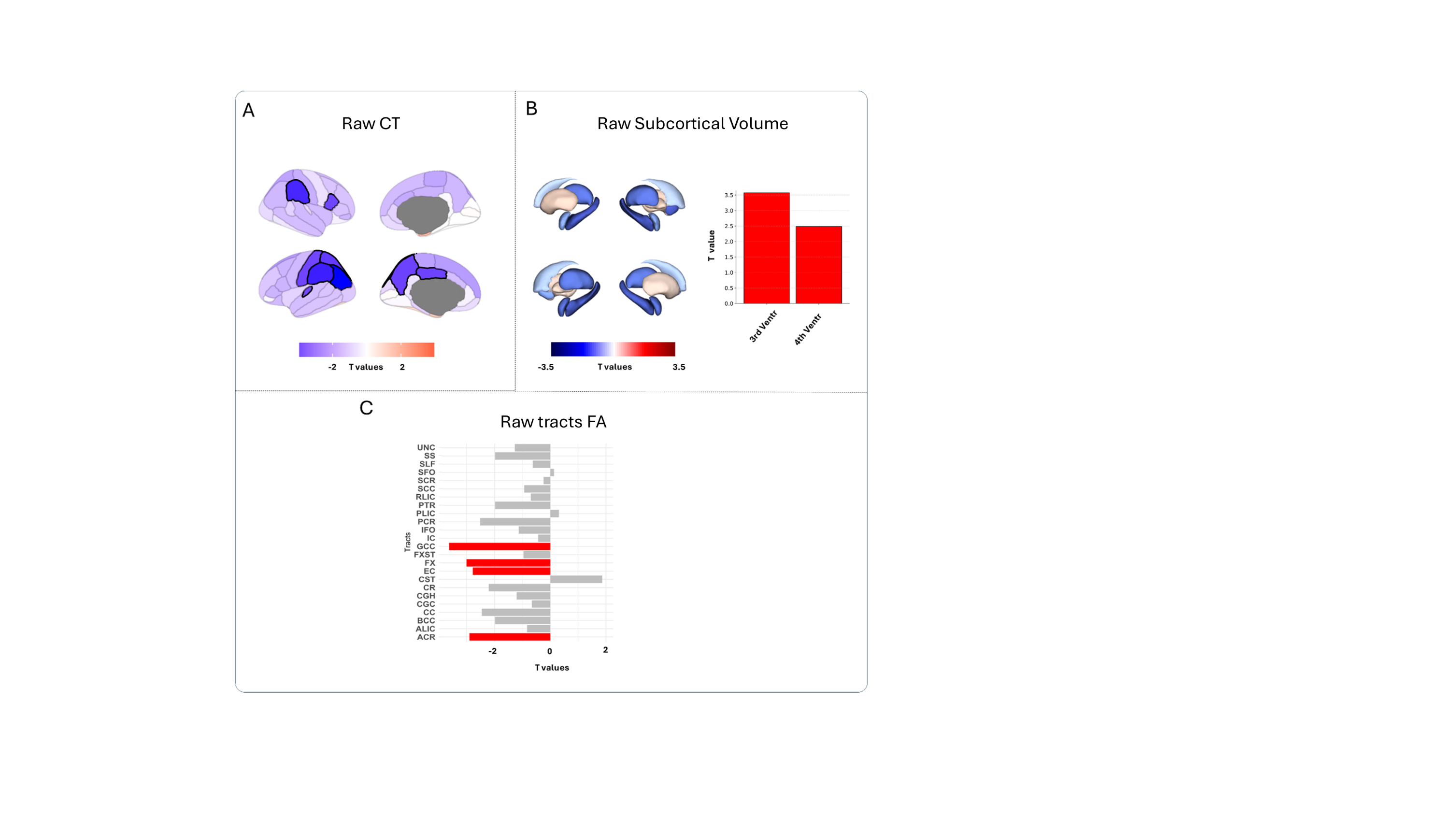

**Fig. S1.** Effect sizes (T-values) obtained using raw structural data , reflecting the contribution of SSD diagnosis, after considering sex, age, estimated Total Intracranial Volume (eTIV), across four structural features: (A) Raw CT , (B) Raw subcortical volumes and (C) Raw FA tracts .

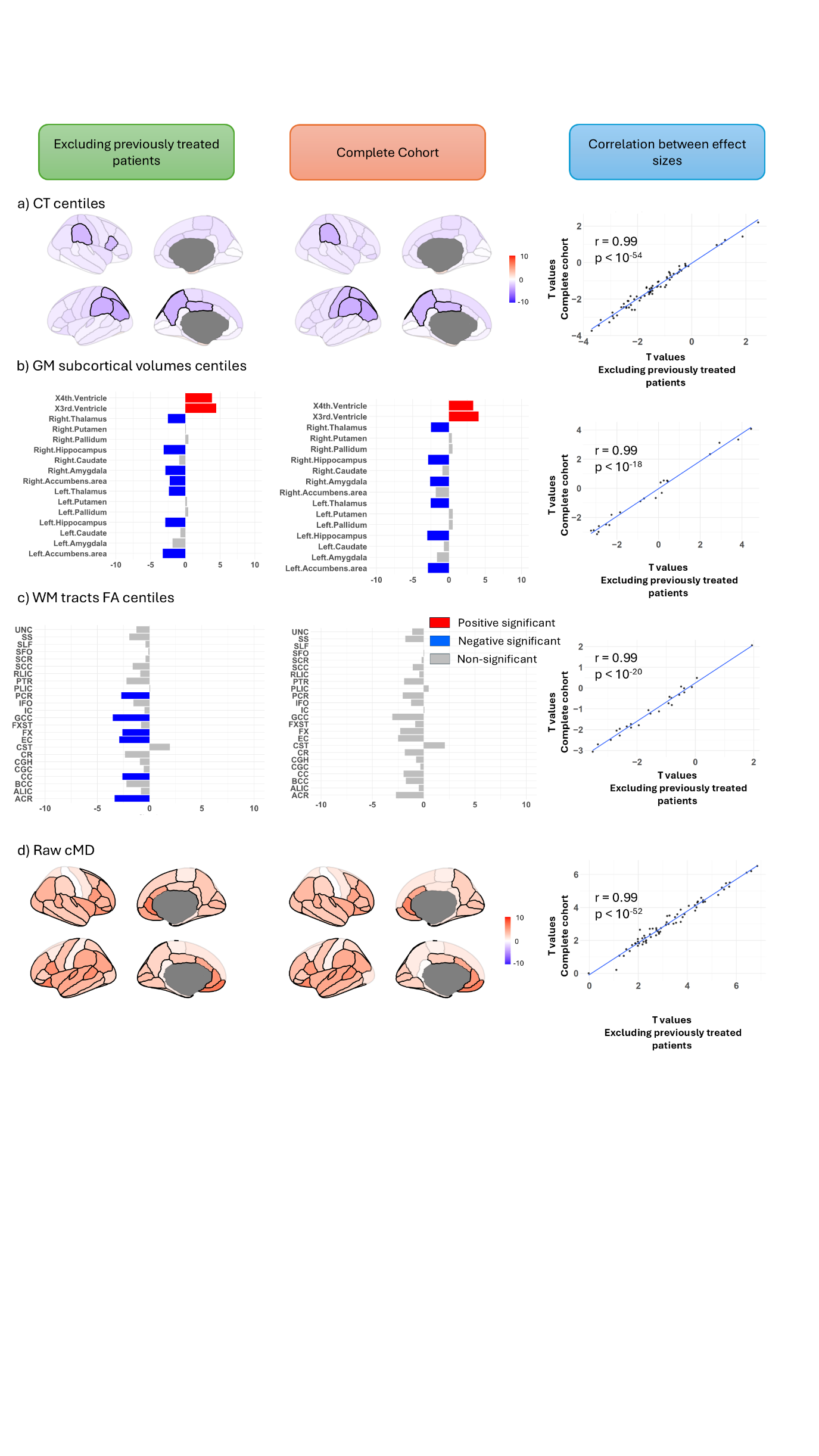

**Fig. S2.** Sensitivity analyses excluding SSD participants with prior antipsychotic exposure. The first column shows effect sizes (T-values) obtained after excluding previously medicated participants, whereas the second column shows the corresponding results from the full cohort analyses. The third column displays the Pearson correlation between the original and sensitivity analyses across regions/features. Structural measures include: (A) CT centiles, (B) subcortical volumetric centiles, (C) FA tract centiles, and (D) raw cMD. Colour scales range from blue (negative T-values indicating reduced structural features in SSD compared to control) to red (positive T-values indicating increased structural features in SSD compared to control). Regions that did not survive FDR correction are shown with desaturated semi-transparent shading, whereas significant regions are displayed in solid colours. Significant tracts and ventricles are shown in solid red.

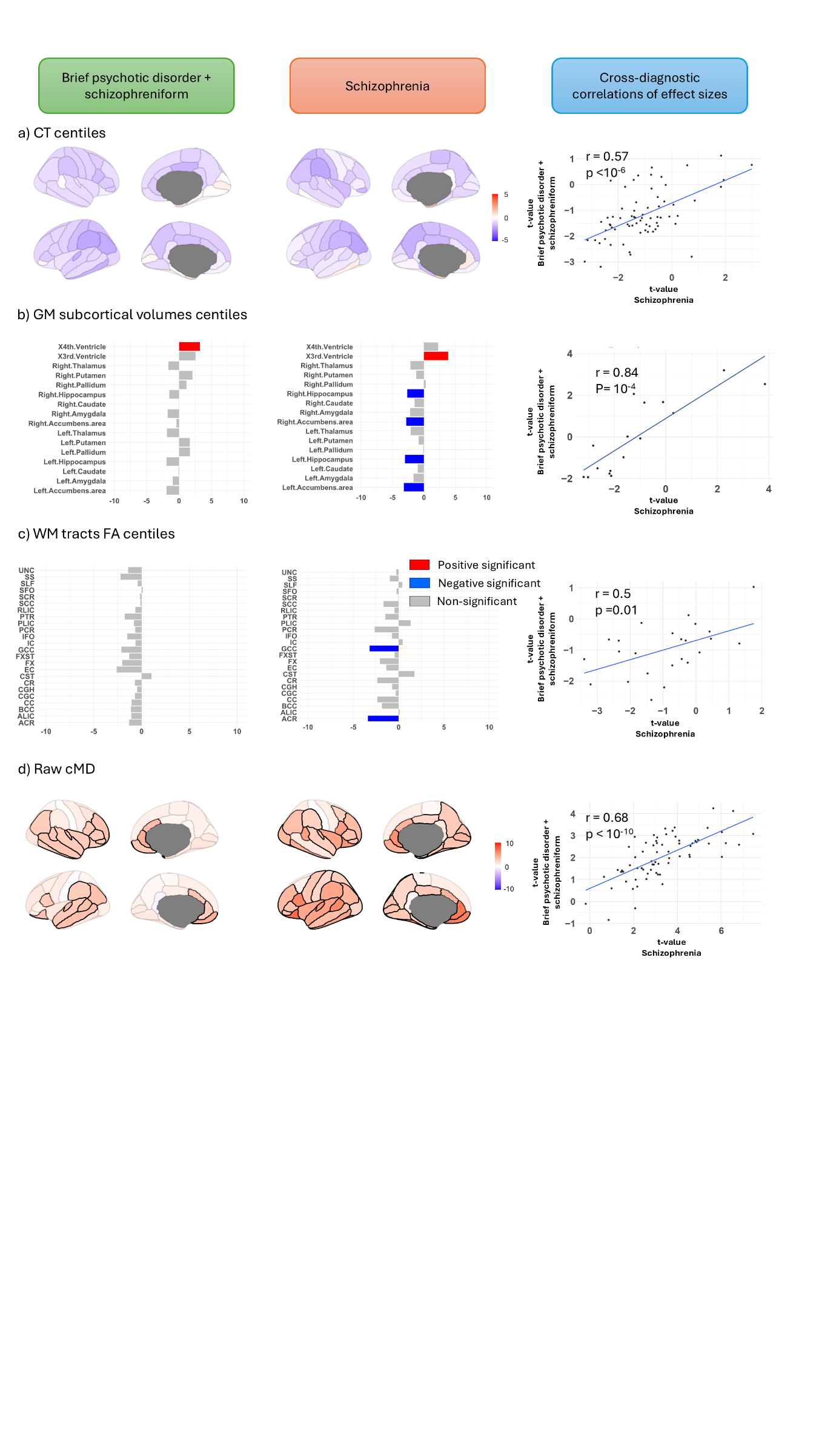

**Fig. S3.** Sensitivity analyses assessing the influence of diagnosis. The first column shows effect sizes (T-values) obtained when restricting the SSD cohort to individuals with brief psychotic disorder or schizophreniform disorder, whereas the second column shows the corresponding results obtained for individuals with schizophrenia. The third column displays the Pearson correlation between effect sizes across regions. Structural measures include: (A) CT centiles, (B) subcortical volumetric centiles, (C) FA tract centiles, and (D) raw cMD. Colour scales range from blue (negative T-values indicating reduced structural features in SSD compared to control) to red (positive T-values indicating increased structural features in SSD compared to control). Regions that did not survive FDR correction are shown with desaturated semi-transparent shading, whereas significant regions are displayed in solid colours. Significant tracts and ventricles are shown in solid red.

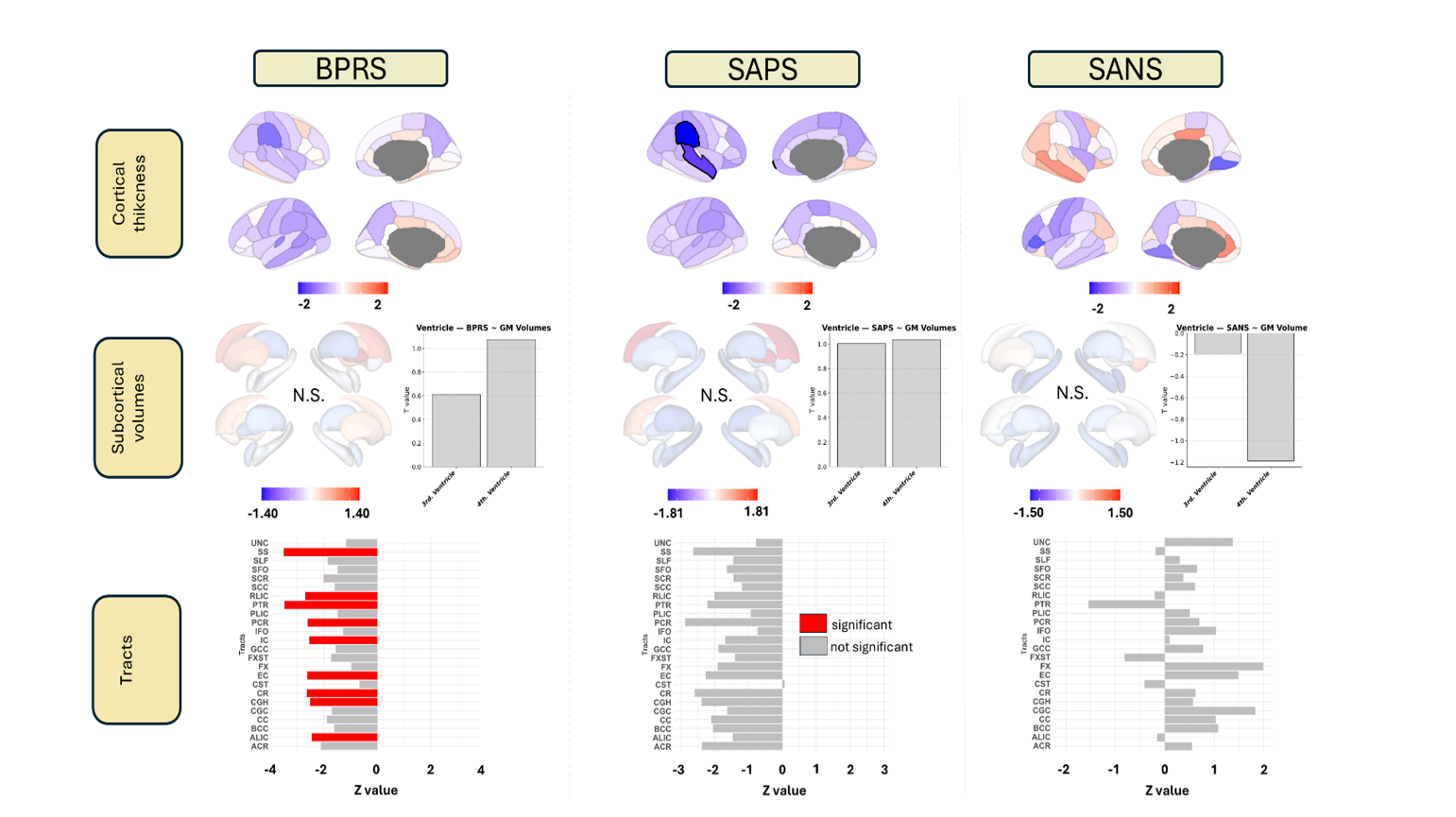

**Fig. S4.** Effect sizes (T-values) showing associations between symptom scores (BPRS, SAPS, SANS) and structural features (raw regional CT, raw subcortical volumes and raw FA tract) using raw structural data.
